# MDA5-autoimmunity and Interstitial Pneumonitis Contemporaneous with the COVID-19 Pandemic (MIP-C)

**DOI:** 10.1101/2023.11.03.23297727

**Authors:** Khizer Iqbal, Saptarshi Sinha, Paula David, Gabriele De Marco, Sahar Taheri, Ella McLaren, Sheetal Maisuria, Gururaj Arumugakani, Zoe Ash, Catrin Buckley, Lauren Coles, Chamila Hettiarachchi, Gayle Smithson, Maria Slade, Rahul Shah, Helena Marzo-Ortega, Mansoor Keen, Catherine Lawson, Joanna Mclorinan, Sharmin Nizam, Hanu Reddy, Omer Sharif, Shabina Sultan, Gui Tran, Mark Wood, Samuel Wood, Pradipta Ghosh, Dennis McGonagle

**Author notes:** **Corresponding Authors**: Dennis McGonagle PhD FRCPI, Leeds Teaching Hospitals NHS Trust, Rheumatology Department, Leeds, United Kingdom | University of Leeds, Leeds Institute of Rheumatic and Musculoskeletal Medicine, Leeds, United Kingdom |; Pradipta Ghosh, M.D., Professor, Departments of Medicine and Cellular and Molecular Medicine, University of California San Diego; 9500 Gilman Drive (MC 0651), George E. Palade Bldg, Rm 232, 239; La Jolla, CA 92093, USA | |. These authors contributed Equally as first author (Khizer Iqbal, Saptarshi Sinha, Paula David).

## Abstract

**Background:** Anti-MDA5 (Melanoma differentiation-associated protein-5) positive dermatomyositis (MDA5^+^-DM) is characterised by rapidly progressive interstitial lung disease (ILD) and high mortality. MDA5 senses single-stranded RNA and is a key pattern recognition receptor for the SARS-CoV-2 virus.

**Methods:** This is a retrospective observational study of a surge in MDA5 autoimmunity, as determined using a 15 muscle-specific autoantibodies (MSAs) panel, between Janurary 2018-December 2022 in Yorkshire, UK. MDA5-positivity was correlated with clinical features and outcome, and regional SARS-CoV-2 positivity and vaccination rates. Gene expression patterns in COVID-19 were compared with autoimmune lung disease and idiopathic pulmonary fibrosis (IPF) to gain clues into the genesis of the observed MDA5^+^-DM outbreak.

**Results:** Sixty new anti-MDA5+, but not other MSAs surged between 2020-2022, increasing from 0.4% in 2019 to 2.1% (2020), 4.8% (2021) and 1.7% (2022). Few (8/60) had a prior history of confirmed COVID-19, peak rates overlapped with regional SARS-COV-2 community positivity rates in 2021, and 58% (35/60) had received anti-SARS-CoV-2 RNA vaccines. Few (8/60) had a prior history of COVID-19, whereas 58% (35/60) had received anti-SARS-CoV-2 RNA vaccines. 25/60 cases developed ILD which rapidly progression with death in 8 cases. Among the 35/60 non-ILD cases, 14 had myositis, 17 Raynaud phenomena and 10 had dermatomyositis spectrum rashes. Transcriptomic studies showed strong *IFIH1* (gene encoding for MDA5) induction in COVID-19 and autoimmune-ILD, but not IPF, and *IFIH1* strongly correlated with an IL-15-centric type-1 interferon response and an activated CD8+ T cell signature that is an immunologic hallmark of progressive ILD in the setting of systemic autoimmune rheumatic diseases. The *IFIH1* rs1990760TT variant blunted such response.

**Conclusions:** A distinct pattern of MDA5-autoimmunity cases surged contemporaneously with circulation of the SARS-COV-2 virus during COVID-19. Bioinformatic insights suggest a shared immunopathology with known autoimmune lung disease mechanisms.

## Introduction

Dermatomyositis (DM) is a systemic autoimmune disease characterized by muscle and skin inflammation and potentially fatal-internal organ involvement, typically interstitial lung disease (ILD) leading to progressive pulmonary fibrosis. The first autoantibody defined in DM was anti-Jo-1, which targets the enzyme histidyl-tRNA synthetase. Since then, many muscle-specific autoantibodies (MSA) emerged, often linked to different clinical phenotype patterns and different MHC-II associations that further underpin the veracity of the autoimmunity concept in DM ^1–4^.

One of the well-recognised clinical phenotype of DM is clinically amyopathic dermatomyositis (CADM) that is associated with rapidly progressive ILD and is attributed to the Retinoic acid-inducible gene 1 (RIG-1)-like receptor family gene, *IFIH1*, which encodes the protein Melanoma differentiation-associated protein-5 (MDA5) ^5^. Most MDA5+ cases predating the COVID-19 pandemic reported significant ILD but a relative lack of myositis or the classical DM heliotropic rash; instead, they showed cutaneous phenotypes including skin ulceration and tender palmar papules ^6^.

Here we report a surge in the rate of anti-MDA5 positivity testing in our region (Yorkshire) in the second year of the COVID-19 pandemic, which was notable because this entity is relatively rare in the UK. This was intriguing because MDA5 is a RIG-1 helicase ^7^ tasked to sense single-stranded RNA and is a key pattern recognition receptor for the contemporary SARS-CoV-2 virus ^8^. Variants of the MDA5 protein-coding gene, *IFIH1* (rs1990760 TT) have recently been shown to confer protection in COVID-19 infections and experienced better outcomes ^9^.

In this retrospective study, we explored the phenotypes and epidemiological factors associated with the cluster of MDA5^+^-related disease at our centre which provides autoantibody testing for a 3.6 million-large population (**Figure 1-***Steps 1-2*). We describe this phenomenon as *<u>M</u>*DA5 autoimmunity with *i*nterstitial *<u>p</u>*neumonitis cotemporaneous with the *<u>C</u>*OVID-19 pandemic (MIP-C) that reflects the different epidemiology and clinical patterns reported herein compared to previously defined MDA5 related autoimmunity. We also leveraged transcriptomic datasets to explore putative mechanisms of this emergent MDA5-associated disease in the setting of SARS-CoV-2 infection (**Figure 1-***Step 3*). Specifically, as post COVID pneumonia is associated with pulmonary fibrosis, we leveraged datasets to compare acute COVID-19 lung disease, autoimmune lung disease and idiopathic pulmonary fibrosis (IPF) to gain clues into the genesis of the observed MDA5^+^-DM outbreak. Finally, we presented a working model that links severity of anti-viral cytokine response to *IFIH1* induction and genetics and ultimately, to the distinct immunophenotype specific for MSA-associated progressive ILD (**Figure 1-** *Step 4*). These findings provide insights into the observed surge in anti-MDA5 positivity during the COVID-19 pandemic and the potential role of RNA viruses in rapidly progressive ILD and other autoimmune conditions.

**Figure 1.**
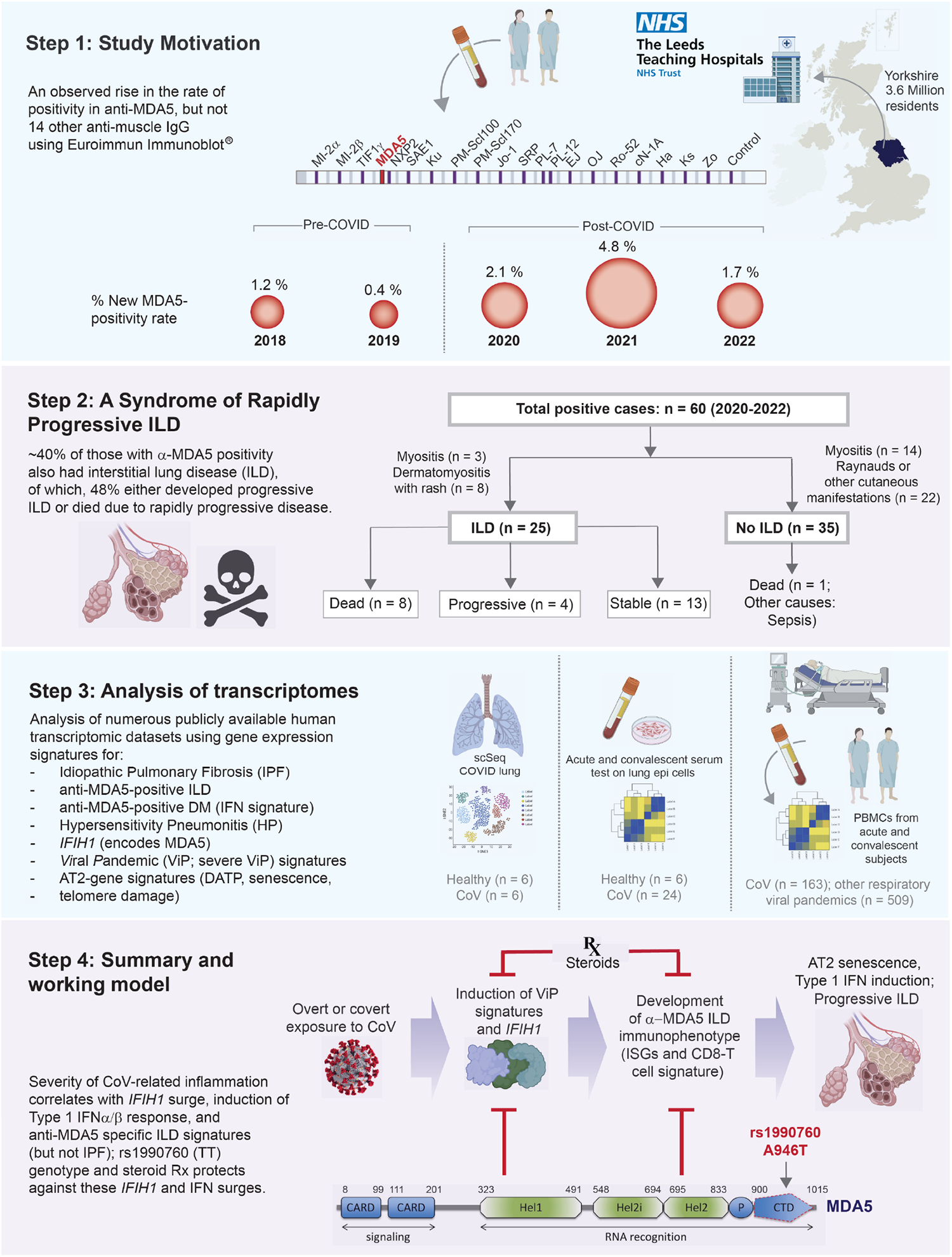
Study Motivation, Design and Major Findings: Schematic summarizes the retrospective study design and motivation (Step 1), and the phenotypical and epidemiological features observed in our cohort (Step 2). It also highlights the analyses of diverse transcriptomics datasets (Step 3) which were carried out to interrogate how COVID-19 infection interacts with IFIH1 gene (encodes MDA5) and disease risk signatures for the development of interstitial pneumonitis of various types. Finally, we summarize findings and propose a working model linking epidemiologic findings to the insights drawn from transcriptomic analyses.

## Methods

### Study design

The Leeds Teaching Hospitals NHS Trust serves as an immunology laboratory reference for the wider Yorkshire region of the UK. We audited the increased anti-MDA5 positivity in relationship to other MSA (Euroimmun immunoblot©) that included MDA5^+^ cases. This was based on both increased rate of anti-MDA5 related immunology reporting and multiple physicians seeing MDA5 related disease for the first time, combined with emergent literature reporting COVID-19 era anti-MDA5-related disease ^1–4,10–28^. We collected data on the number of MDA5+ tests per year between January 2018 to December 2022. The clinical notes review focused on patterns of symptomatic MDA5 disease (including degree of ILD); muscle or other organs involvement, therapy, therapy responses and survival data.

We also leveraged Public Health England (PHE) data on SARS-CoV-2 monthly positivity rates in the Yorkshire region. We also evaluated data on lung involvement and concomitant SARS-CoV-2 infection, recent SARS-CoV-2 infection or recent SARS-CoV-2 vaccination or both infection and vaccination by searching for confirmed PCR positivity for infection or confirmation of vaccination status including number of vaccines administered as gleaned from “NHS spine” system, a system that supports the IT infrastructure for health and social care for England, joining together over 44,000 healthcare systems in 26,000 organizations ^29^.

### Ethics Statement

Ethics committee/ Institutional Research Board (IRB) of University of Leeds, UK, waived ethical approval for this work. This study was reported according to the “CAse REports” (CARE) guidelines [https://www.care-statement.org/]. All participants recruited granted verbal or written consent to the local treating physicians for the use of their anonymized data. An approved retrospective audit of service delivery at our institution, and a formal IRB approval was not needed.

### Computational Analyses

#### Transcriptomic Datasets and Data Analyses

To explore potential mechanistic links between COVID infection and lung disease we analyzed several publicly available datasets (COVID-19, n = 240; ILD, n = 316; viral pneumonitis, n = 1038), a complete catalog of which is presented in **Supplemental Information 1**). To decipher which immunophenotype is induced in the setting of COVID-19, previously validated lung or PBMC-based gene signatures from distinct lung diseases were used: (i) idiopathic pulmonary fibrosis (IPF); (ii) hypersensitivity pneumonitis (HP); (iii) systemic autoimmune rheumatoid diseases (SARDs) such as systemic sclerosis and MDA5^+^-DM; and (iv) well-defined signatures of so called “AT2 cytopathies”, i.e., ER stress, stem cell dysfunction, senescence, and telomere shortening, which have been implicated in driving fibrotic lung disease after diffuse alveolar injury, as in the setting of severe COVID-19 ^30^ and IPF ^31^). All gene signatures used in this work are presented in an excel sheet, alongside the original source articles (**Supplemental Information 2**).

#### Single Cell RNA Sequencing Analysis

Single Cell RNASeq data from GSE145926 was downloaded from Gene Expression Omnibus (GEO) in the HDF5 Feature Barcode Matrix Format. The filtered barcode data matrix was processed using Seurat v3 R package. B cells (CD19, MS4A1, CD79A), T cells (CD3D, CD3E, CD3G), CD4 T cells (CCR7, CD4, IL7R, FOXP3, IL2RA), CD8 T cells (CD8A, CD8B), Natural killer cells (KLRF1), Macrophages, Monocytes and DCs (TYROBP, FCER1G), Epithelial (SFTPA1, SFTPB, AGER, AQP4, SFTPC, SCGB3A2, KRT5, CYP2F1, CCDC153, TPPP3) cells were identified using relevant gene markers using SCINA algorithm.

Several publicly available microarrays and RNASeq databases were downloaded from the National Center for Biotechnology Information (NCBI) Gene Expression Omnibus (GEO) server. Gene expression summarization was performed by normalizing Affymetrix platforms by RMA (Robust Multichip Average) and RNASeq platforms by computing TPM (Transcripts Per Millions) values whenever normalized data were not available in GEO. We used log2(TPM +1) as the final gene expression value for analyses. GEO accession numbers are reported in figures and text. A catalog of all datasets analyzed in this work can be found in **Supplemental Information 1**.

#### Gene Expression Analyses

The expression levels of all genes in these datasets were converted to binary values (high or low) using the *StepMiner* algorithm ^32,33^ which undergoes an adaptive regression scheme to verify the best possible up and down steps based on sum-of-square errors. The steps are placed between data points at the sharpest change between expression levels, which gives us the information about threshold of the gene expression-switching event. To fit a step function, the algorithm evaluates all possible steps for each position and computes the average of the values on both sides of a step for the constant segments. An adaptive regression scheme is used that chooses the step positions that minimize the square error with the fitted data. Finally, a regression test statistic is computed as follows:

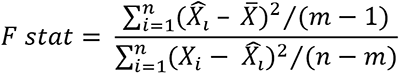

Where *X*_i_ for *i* = 1 to *n* are the values, 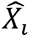 for *i* = 1 to *n* are fitted values. M is the degrees of freedom used for the adaptive regression analysis. *X̄* is the average of all the values:

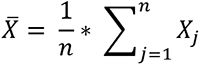

For a step position at k, the fitted values 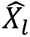 are computed by using

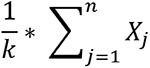

for *i* = 1 to *k* and

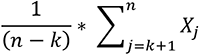

> for *i* = *k* + 1 to *n*.

Gene expression values were normalized according to a modified Z-score approach centered around *StepMiner* threshold (formula = (expr – SThr)/3*stddev). The normalized expression values for every genes were added together to create the final score for the gene signature. The samples were ordered based on the final signature score. Classification of sample categories using this ordering is measured by ROC-AUC (Receiver Operating Characteristics Area Under The Curve) values. Welch’s Two Sample t-test (unpaired, unequal variance (equal_var=False), and unequal sample size) parameters were used to compare the differential signature score in different sample categories. Violin plots are created using python seaborn package version 0.10.1. Differentially expressed genes are identified using DESeq2 package in R.

#### Correlation plot

StepMiner normalized composite score of gene signatures were plotted against each other for all the patients. For each two signatures, linear least-squares regression has been obtained using SciPy LLS model (scipy.stats.linregress). R^2^ and p-value for each pair of signatures are plotted as heatmap using seaborn (seaborn.heatmap) package.

#### Multivariate Analyses

To assess which factor(s) may influence MDA5 induction upon exposure to SARS-CoV2, multivariate regression has been performed on the bulk sequence COVID-19 PBMC datasets (GSE233626 [updated with additional variables from GSE168400] and GSE233627 (updated with additional variables from GSE177025). Multivariate analysis of GSE233626 models the degree of *IFIH1* induction in samples (base variable) as a linear combination of gender, age, ventilation, hypoxemia with/without genotype. Multivariate analysis of GSE233627 also models the degree of *IFIH1* induction in samples (base variable) as a linear combination of the same variables as above, and an additional variable-that of treatment with systemic corticosteroids. Here, the statsmodels module from python has been used to perform Ordinary least-squares (OLS) regression analysis of each of the variables. The choice of these datasets was driven by the criteria that they are high quality datasets with maximal unique patient samples. The p-value for each term tests the null hypothesis that the coefficient is equal to zero (no effect).

#### Data and Code Availability

All codes and datasets used in this work can be found at https://github.com/sinha7290/COVID_mda5.

## Results

### MDA5 positivity between 2018-2022

Between January 2018 and December 2019, 6 new MDA5^+^ cases were identified, representing 1.2% and 0.4% MSA immunoblot positivity in the respective years (**Figure 2A**). However, commencing in 2021, after the second UK SARS-CoV-2 infection wave, we noted an increase in new MDA5^+^ cases (**Figure 2**). The total numbers of new cases were 9, 35 and 16 in 2020, 2021 and 2022 respectively (**Figure 2A**). Irrespective of the fact that MSA requisitions requests approximately doubled during the same period of time, an increased rate of MDA5 positivity was evident, rising from 1.2% in 2018 and 0.4% in 2019 to to 2.2% in 2020, 4.8% in 2021 and decreasing to 1.7% in 2022. The other MSAs did not exhibit this striking pattern of increase (**Figure 2A**-*top*).

**Figure 2.**
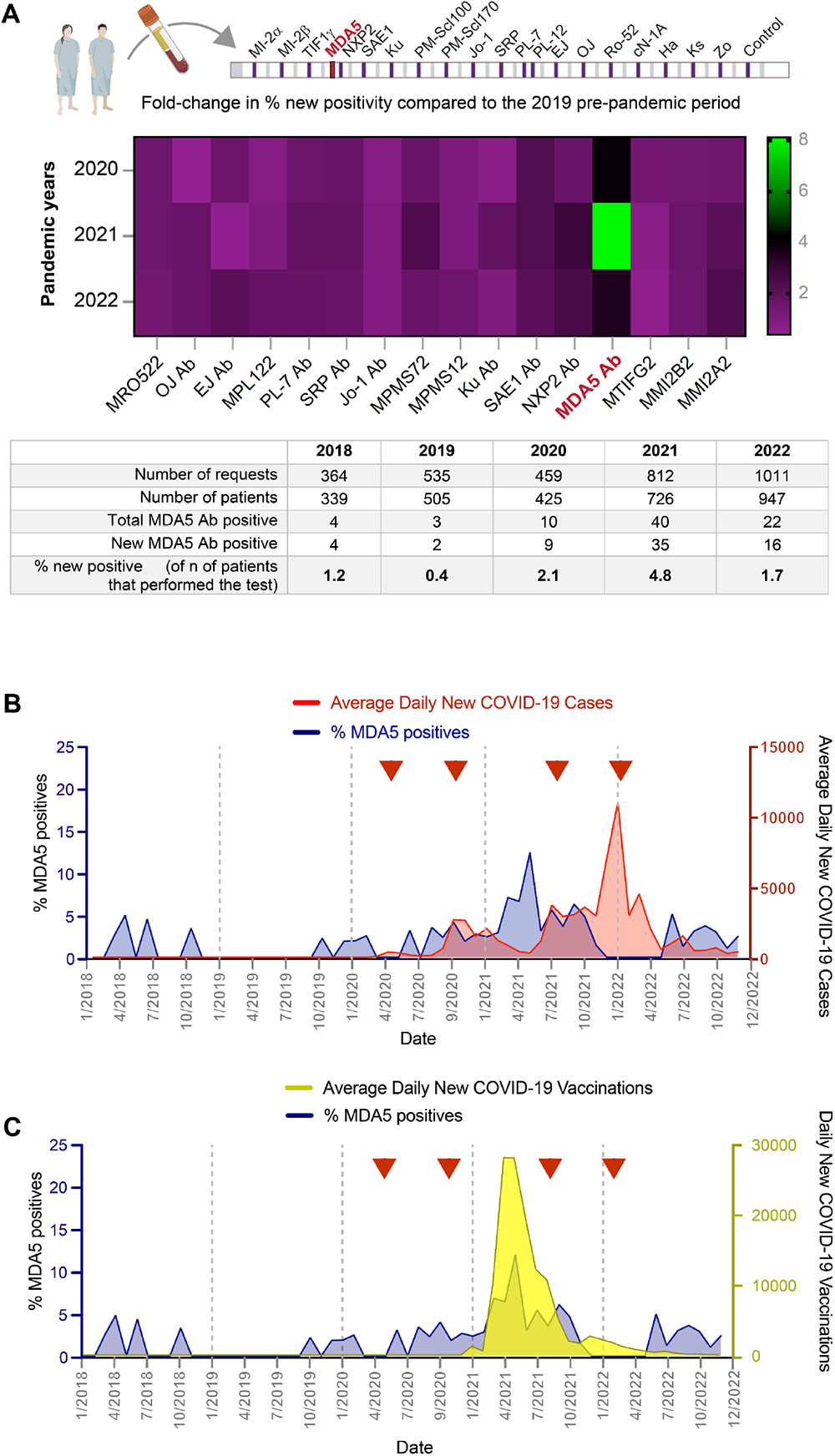
–Rate of MDA5+ testing 2018 to 2022. **A.** Heatmap (top) shows the fold change in MDA5+ for each of the tested muscle-specific autoantibodies (MSAs), including anti-MDA5 (using Euroimmun immunoblot©). Table (bottom) provides the actual patient numbers. **B-C**. Graphs display the overlay of newly detected anti-MDA5 positivity (blue; A-B) with either total COVID-19 cases (red; A) or the rate of new vaccination (yellow; B) that were reported in the Yorkshire and Humber regions since Jan 2021 to Dec 2022. The COVID-19 case rates and vaccination rates were obtained from the UK.gov database (https://coronavirus.data.gov.uk/). Red arrowheads denote the four waves of COVID-19 cases.

### Clinical features of the 60 new MDA5 positive cases

Thirty-two/60 were of white ethnic background [either British or other still classified as white, according to 2021 UK census methodology ^34^]. Three/60 were of Asian/Asian British (all of these Indian/Pakistani) background; 2 were of Black Caribbean and 1 of Black African ethnic background and 4 were considered “any other ethnic group”. Four patient was of other Asian background (not Chinese) with no ethnicity data for 14/60 patients.

All 60 patients experienced some features consistent with an autoimmune disease, their average age was 56.17 years (median 56; standard deviation 19.9; absolute range 9-90; inter-quartile range 43.75-71.25) and 36/60 (60%) were female. Of the 60 patients, 25 developed ILD with a mean age 60.28 years; median 66; standard deviation 18.56; absolute range 12-90; and inter-quartile range 51-73. Twelve/25 (48%) were females. Almost half of this subgroup (12/25, 48%) rapidly progressed and 8 of them died. By contrast, just 1 fatality was observed in the 35 patients who did not develop ILD (sepsis-related). Out of 4 new paediatric patients in this series, none were fatal and none were vaccinated against SARS-CoV2.

The 35 patient non-ILD group had a mean age of 53.23 years (median 54; standard deviation 20.6; absolute range 9-89; inter-quartile range 40-69). 24/35 (68.6%) were females; 4/60 were < 18 years old. Although the non-ILD subgroup was younger than their ILD counterparts (**Table 1**), this difference was not statistically significant (Student T test p-value = 0.179). The two subgroups did not differ in terms of gender representation (Fisher’s exact test p-value 0.120).

**Table 1:**
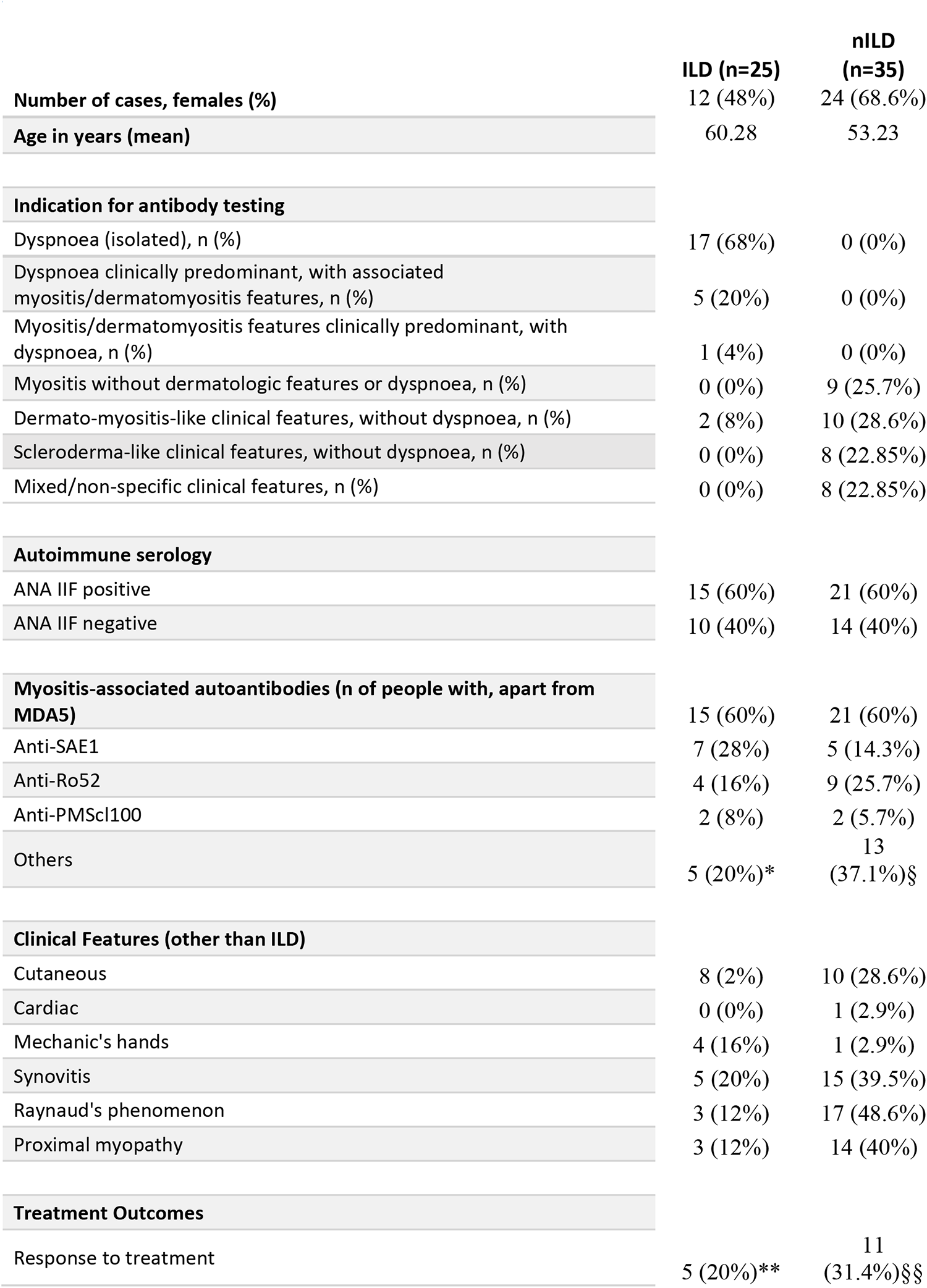
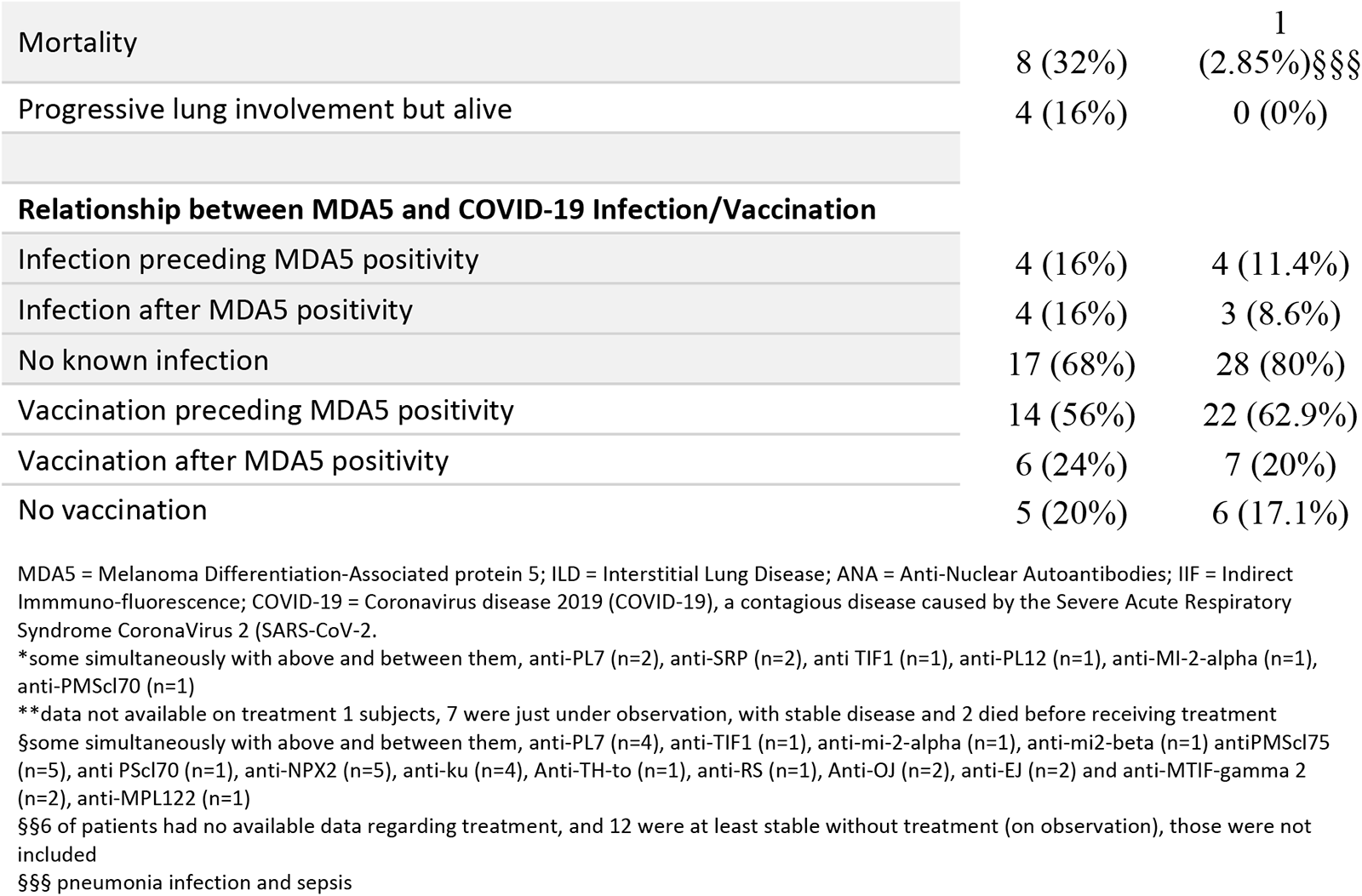
MDA5+ Disease split up into ILD and non ILD cases.

The main indication for requesting MSA testing in the ILD subgroup was dyspnoea with and without associated myositic/DM features (**Table 1**, and **Supplemental Table 1**). The indication for performing such testing in the non-ILD subgroup was cutaneous manifestations of DM or scleroderma-like clinical features, as well as proximal myopathy (**Table 1** and **Supplemental Table 2**). There was one case of confirmed myocarditis. The creatine kinase (CK) at baseline was available for 50/60 patients and its average was 811.78 units per liter (U/L), however, the median was 90.5 U/L in keeping with CADM phenotype (IQR 56.75-199); there was no statistically significant difference between ILD and non-ILD groups (median 78 vs. 115, respectively Mann-whitney U test p value of 0.186). Of 35 non-ILD cases, at least 9 (missing data on imaging for 9/35 patients) had muscle MRI, of them 5 were compatible with myositis. Details of therapy are shown for each case in **Supplemental Tables 1–2**.

### MDA5 positive ILD outcomes

As expected the prognosis was poorer in the 25 patients in the ILD patients. Chest CT was available in 24/25 cases, which demonstrated fibrosis and associated ground glass changes in 6/25 cases; fibrotic changes only in 8/25 cases; ground glass changes only in 9/25 cases; ground glass changes with pneumomediastinum in 1 case. In keeping with the MDA5 phenotype, 8/25 patients progressed, most rapidly, and died despite intensive therapy; 4/25 developed progressive lung disease; 12/25 stabilised with or without specific therapy. There is one patient with no available data regarding response to treatment. There was no evidence of myocarditis in this subset and mortality was due to pulmonary disease (**Supplemental Table 1**). The only patient of paediatric age in this group remains stable.

### Non-ILD MDA5 positive disease

All MDA5^+^ cases had some clinical features of autoimmune connective tissue disease, including cutaneous manifestations of DM or Raynaud’s phenomenon (**Table 1** and **Supplemental Table 2**). More patients in the non-ILD subgroup developed cutaneous rash (10/35) and Raynaud’s phenomenon (17/35), sometimes both, and proximal myopathy (14/35) with only 1/35 developing “mechanic’s hands” (**Supplemental Table 2**).

### Autoantibody testing

There was no difference in ANA positivity between the ILD subgroup and the the non ILD subgroup (60% positive in both groups, as determined by immunofluorescence). In both subgroups SAE1 and Ro-52 were the auto-antibodies most often positive concomitantly to the anti-MDA5. 15/25 patients in the ILD subgroup had additional MSA antibodies as compared to 21/35 in the non-ILD subgroup (χ^2^ test p-value = 0.930). 4/8 (50%) of patients who died in ILD subgroup had additional MSA antibodies, being anti-small ubiquitin-like modifier-1 (SAE-1) MSA the most common, evident in 3/4.

### Relationship to COVID-19 infection or vaccination

In lieu of patient autoimmune symptoms and signs, MDA5+ testing emerged only after the second and third SARS-CoV-2 wave in the Yorkshire region (**Figure 2B**). Also, the highest rate of MDA5 positivity did occur during higher community SARS-COV-2 positivity during 2021 but the highest rate of SARS-CoV-2 circulaiton was not followed by an immediate increased MDA5+ testing (**Figure 2B**). 8/60 had confirmed COVID-19 before anti-MDA-5+ test performed, and 7/60 were infected after the diagnosis, with 2 of them flaring during the infection. Overall, 15/60 had confirmed SARS-CoV-2 infecton with only 8/25 positive in the ILD subgroup and 7/35 in the non ILD subgroup.

As for vaccinations, the overall uptake of SARS-CoV-2 vaccination in the UK and Yorkshire region was 90% and we saw a strong overlap between vaccination timing in 2021 and the surge in MDA5+ disease (**Figure 2C**) but such a close link with monthly confirmed infections was lacking (**Figure 2B-C**). 49/60 (81.6%) cases had documented evidence of SARS-CoV-2 vaccination; 20/25 in the ILD subgroup and 29/35 in non-ILD subgroup. 36/60 (60%) cases were vaccinated before anti-MDA5 positivity, 14/60 were vaccinated after, of which 2/14 had a disease flare. 11/60 (5/25 ILD and 6/35 non-ILD) were not vaccinated at any point. In the ILD group, 14/25 (56%) were vaccinated preceding the MDA5+ test, while in the nILD group 22/35 (62.9%) (χ^2^ test p-value = 0.271).

Accordingly, most of the MDA5^+^ cases had either confirmed infection or confirmed SARS-CoV-2 vaccination. All the 4 patients of paediatric age, were not vaccinated (all of these developed MDA5 positivity after the pandemic started). Time-relationship to vaccine and infection for each individual is summarized in **Supplemental Tables 1-2**.

### COVID-19 lungs show induction of MDA5 (*IFIH1*) gene and signatures of SARD-related ILD

We leveraged available transcriptomic datasets to explore potential mechanisms of MDA5+ disease in the setting of COVID-19. Analysis of bronchoalveolar lavage fluid from COVID-19 lungs by single cell RNA sequencing (scSeq; **Figure 3A**) confirmed that *IFIH1* is induced significantly in diverse cells of the lavage fluid (**Figure 3B**; *arrow, bubble plot*), alongside the robust induction of a set of several previously validated signatures (**Figure 3B**):

i. an intense IL-15-centric type 1 interferon (IFN) response, a.k.a, the *<u>Vi</u>*ral *<u>P</u>*andemic (ViP) and its subset, severe(s)ViP signatures that was identified and rigorously validated using machine learning (on ∼45,000 samples) which capture the ‘invariant’ host response, i.e., the shared fundamental nature of the host immune response induced by all viral pandemics, including COVID-19 ^35^;
ii. a COVID-19 lung signature ^36^;
iii. a set of 3 signatures indicative of alveolar type two (AT2) cytopathies in fibrotic lung disease, i.e., (a) damage associated transient progenitor (DATP) ^37^, a distinct AT2 lineage that is a central feature of idiopathic pulmonary fibrosis (IPF) ^37–39^; (b) AT2-senescence signature ^40^; and (c) Telomerase dysfunction signature, which was derived from aging telomerase knockout (Terc-/-) mice ^41^. Lung epithelial signatures of IPF were also induced (**Figure 3B**), most consistently in the epithelium. However, gene signatures previously reported in ILDs that are related to systemic autoimmune rheumatic diseases (SARDs), [which include systemic sclerosis (SSc), DM, polymyositis (PM), rheumatoid arthritis (RA), primary Sjögren’s syndrome] were induced in a wide variety of cell types (**Figure 3B**).

**Figure 3.**
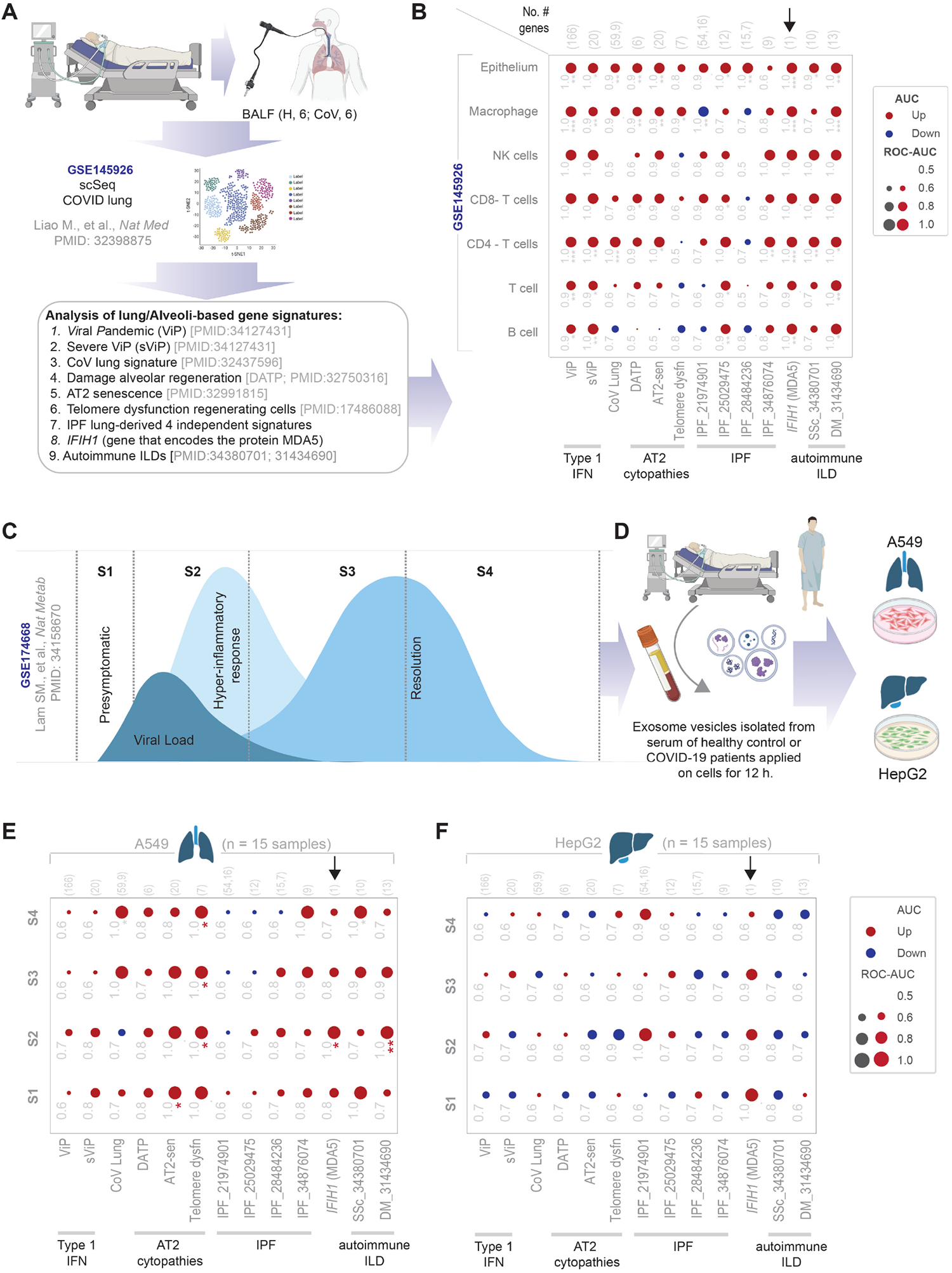
*IFIH1* and autoimmune ILD gene signatures are induced in diverse cell types in CoV lung, including the alveolar epithelium. **A.** Schematic showing the study design for panels A-B. **B**. Bubble plot of ROC-AUC values (radii of circles are based on the ROC-AUC) demonstrating the direction of gene regulation (Up, red; Down, blue) for the classification of various cell types between healthy and CoV lung based on various gene signatures in **Fig 3A**, which includes several signatures of AT2 cytopathies that are encountered and implicated in ILD. Numbers indicate PMIDs. Welch’s two sample (H vs CoV) unpaired t-test is performed on the composite gene signature score (z-score of normalized tpm count) to compute the *p values* [*. P ≤ 0.05 ; **. P ≤ 0.01 ; ***. P ≤ 0.001]. **C-D**. Schematic summarizes the study design for GSE174668. Panel C shows the natural course of COVID-19 which includes pre-symptomatic (S1), hyperinflammatory (S2), resolution (S3) and convalescent (S4) phases. Typically, S1-2 is SARS-CoV-2 RNA positive and has mixed inflammation and immunosuppression as host immune response to the virus. The second half (S3-4) is characterized by host immune response that is geared towards resolution of inflammation and restoration of homeostasis. Exosome-enriched EVs were isolated from fasting plasma from healthy controls and COVID-19 patients from and then applied on two cell types (Panel D) for 12 h at 37°C prior to RNA Seq analysis. **E-F**: Bubble plot of ROC-AUC values (radii of circles are based on the ROC-AUC) demonstrating the direction of gene regulation (Up, red; Down, blue) for the classification of cells treated with EVs from healthy controls vs those isolated from the indicated phase of CoV infection (S1-4) based on various gene signatures in **Fig 3A**, which includes several signatures of AT2 cytopathies that are encountered and implicated in ILD. Numbers indicate PMIDs. Welch’s two sample (H vs CoV) unpaired t-test is performed on the composite gene signature score (z-score of normalized tpm count) to compute the *p values* [*. P ≤ 0.05 ; **. P ≤ 0.01]. BALF, bronchoalveolar lavage fluid; H, healthy; CoV, COVID-19; AT2, alveolar type 2 pneumocytes; DATP, damage-associated transient progenitors; SSc, Systemic scleroderma; Sen, senescence.

When exosome vesicles isolated form the serum of COVID-19 patients during various phases of the disease were applied to 2D cultures of lung or liver epithelial cells (see **Figure 3C-D**), *IFIH1* (see **Figure 3E-F**; *arrows*) and gene signatures of AT2 cytopathies and autoimmune ILD were induced significantly and specifically in the lung, but not liver cells. Consistent with its role as an innate immune sensor of RNA viruses, the serum from the disease phase when viral RNA is detectable (S2 phase) triggered a significant induction in *IFIH1* and autoimmune-ILD signature (but not IPF) (**Figure 3C**). We conclude that both *IFIH1* and autoimmune-ILD signatures were induced *in vivo* and *in vitro* upon exposure to viral RNA.

### Expression of MDA5 (*IFIH1*) gene and signatures of autoimmune ILD in COVID-19 PBMCs

The observed induction of *IFIH1* in the immune cells within the lungs warranted a similar analysis of peripheral blood mononuclear cells (PBMCs) from acute and convalescent COVID-19 subjects, using a set of gene signatures that were previously validated in immune cells (enlisted in **Figure 4A**). We prioritized a dataset that also included the information on the *IFIH1* genotype rs1990760 which has recently been shown to impact the degree of inflammatory response and outcomes in COVID-19 ^9^. *IFIH1* induction tightly and positively correlated with type 1 IFNs (**Figure 4B**; ViP), an *ISG15*^+^ CD8^+^ cytotoxic T-cell signature that was found to be associated with risk of progressive ILD in the setting of MDA5 autoimmunity ^42^ (**Figure 4B**; anti-MDA5-ILD) and a distinctive IFN response that is specific for anti-MDA5+ DM (**Figure 4B**; anti-MDA5-DM IFNs). The rs1990760 TT variant that was found to be protective, showed a clear pattern in each comparison tested; two clear groups were observed in each comparison (**Figure 4C**).

**Figure 4.**
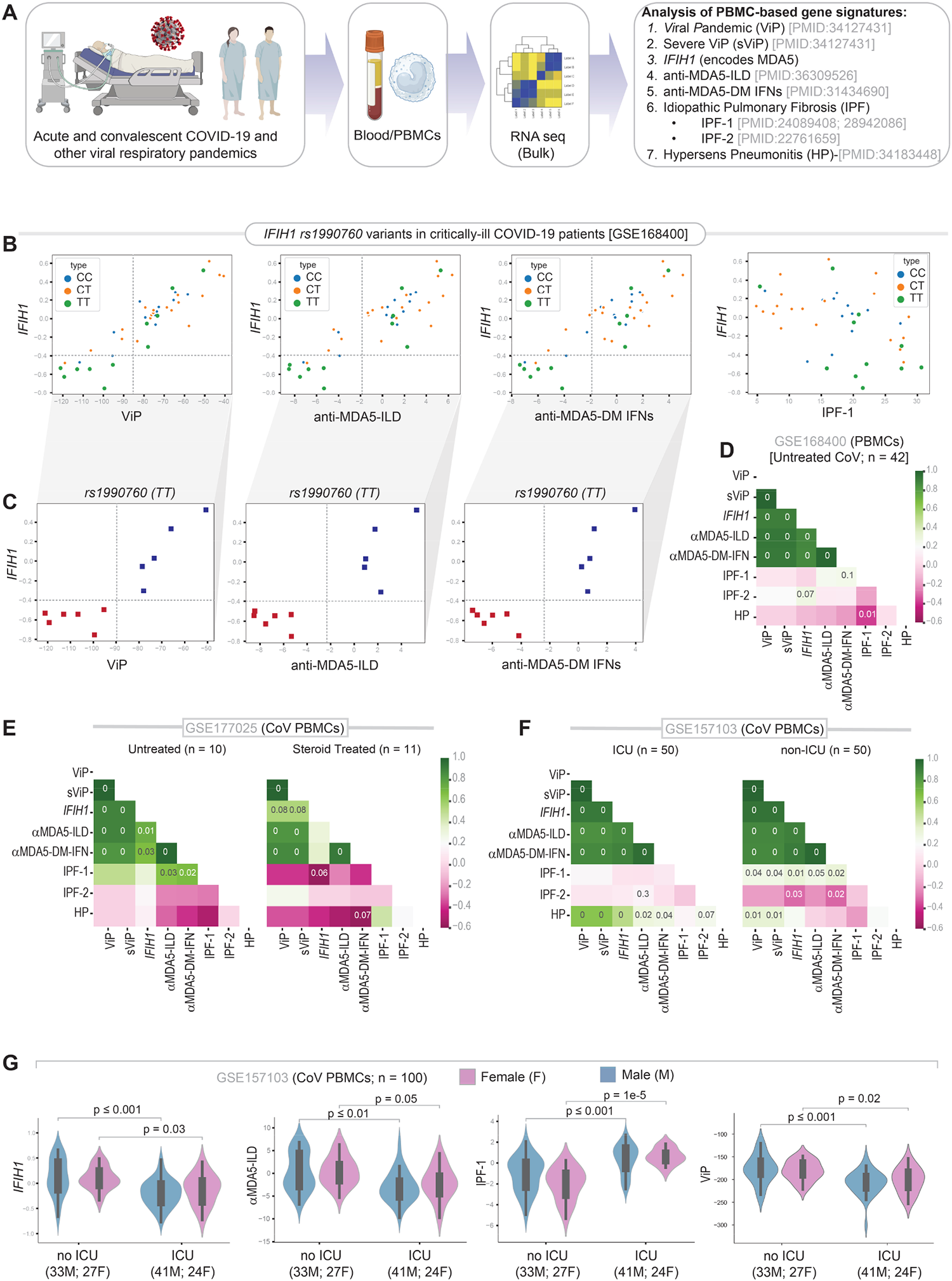
Induction of *IFIH1* in COVID-19 correlates with a Type 1-IFN storm and anti-MDA5-ILD risk signatures in PBMCs. **A.** Schematic of the workflow in this figure, indicating the types of samples analysed and the gene expression signatures tested. **B-C**. Scatter plots show the relationships between *IFIH1* expression (Y axis) and the compositive scores of four different gene expression signatures (X axis) in PBMCs from patients with COVID-19. Top panels in B show all three rs1990760 variant types. Bottom panels in C show just the TT variant. Interrupted lines are drawn arbitrarily to divide the graph into quadrants with low-low and high-high distributions to separate the patients who suppressed *IFIH1* in the TT genotype from those who did not. **D**. Graphical representation of a correlation matrix representing the correlation between the variables in B-C and additional variables, i.e., composite scores of different gene signatures elaborated in panel A. The colour key spans from -1 (magenta) to 1 (green), indicating both strength and direction of correlation. Numbers within the heatmap indicate statistical significance (only significant *p values* are displayed). **E-F**. Correlation matrix showing the correlation between multiple gene signatures (as in D), on two other independent COVID-19 (CoV) patient-derived PBMC datasets. See **Supplementary** Figure 2 for similar analyses on three independent PBMC and whole blood datasets representing other respiratory viral pandemics. **G**. Violin plots show the degree of induction of *IFIH1* (transcripts per million; tpm) and various gene expression signature (composite scores) in male or female patients presenting with moderate (non-ICU) or severe (ICU) COVID-19. Welch’s two sample (ICU vs non-ICU) unpaired t-test is performed on the tpm (for *IFIH1*) or the composite gene signature score (z-score of normalized tpm count) to compute the *p values* (only significant *p values* are displayed).

Unlike autoimmune ILDs, the IPF-related ILDs are known to have a completely distinct immunopathogenesis. We next leveraged a 52-gene PBMC-based IPF signature that was previously discovered ^43^ and subsequently validated as a predictor of IPF progression in a prospective multicenter study ^44^. The expression of *IFIH1* negatively correlated with the 52-gene PBMC-based IPF signature (**Figure 4B**). Negative correlations were observed between *IFIH1* and another independent signature for IPF (IPF-2; **Figure 4D**) and with a signature of hypersensitivity pneumonitis (HP; **Figure 4D**).

All these correlative patterns generally held true when rigorously tested across independent PBMC datasets from diverse patient cohorts, representing COVID-19 (**Figure 4E-F**) and other viral respiratory pandemics (**Supplementary** Figure 1). *IFIH1* induction consistently correlated with a type 1 IFN-centric immune response in MDA5 autoimmunity, but not with the immune response in IPF.

### Impact of severity, gender, steroids and *IFIH1* genotype on MDA5 (*IFIH1*) surge

A subanalysis on the largest PBMC dataset that included information on gender and disease severity revealed that *IFIH1*, anti-MDA5-ILD and ViP signatures were induced in less severe disease which did not warrant ICU-level of care (**Figure 4G**), whereas the 52-gene risk signature for progressive IPF was induced in more severe COVID-19 that required ICU care (**Figure 4G**); these observations held true in both genders.

Next we created a multivariate model to decompose the covariance between the levels of induction of *IFIH1* (base variable), genotype, gender, age, severity of ARDS; as determined using the ratio of PaO2/FiO2) and the need for ventilation (Vent). The *IFIH1* rs1990760 genotype emerged as the strongest determinant of the degree of induction of the *IFIH1*(MDA5) transcript (**Figure 5A***-left*). Age emerged as an independent variable when the rs1990760 TT variant was analyzed independently (**Figure 5A***-middle*); young age was associated with higher levels of induction of *IFIH1* transcripts. Gender and the need for ventilation were covariates when the rs1990760 CT/CC variants were analyzed independently (**Figure 5A***-right*); female gender and moderate disease not requiring ventilator support was associated with a higher level of *IFIH1* transcript surge.

**Figure 5.**
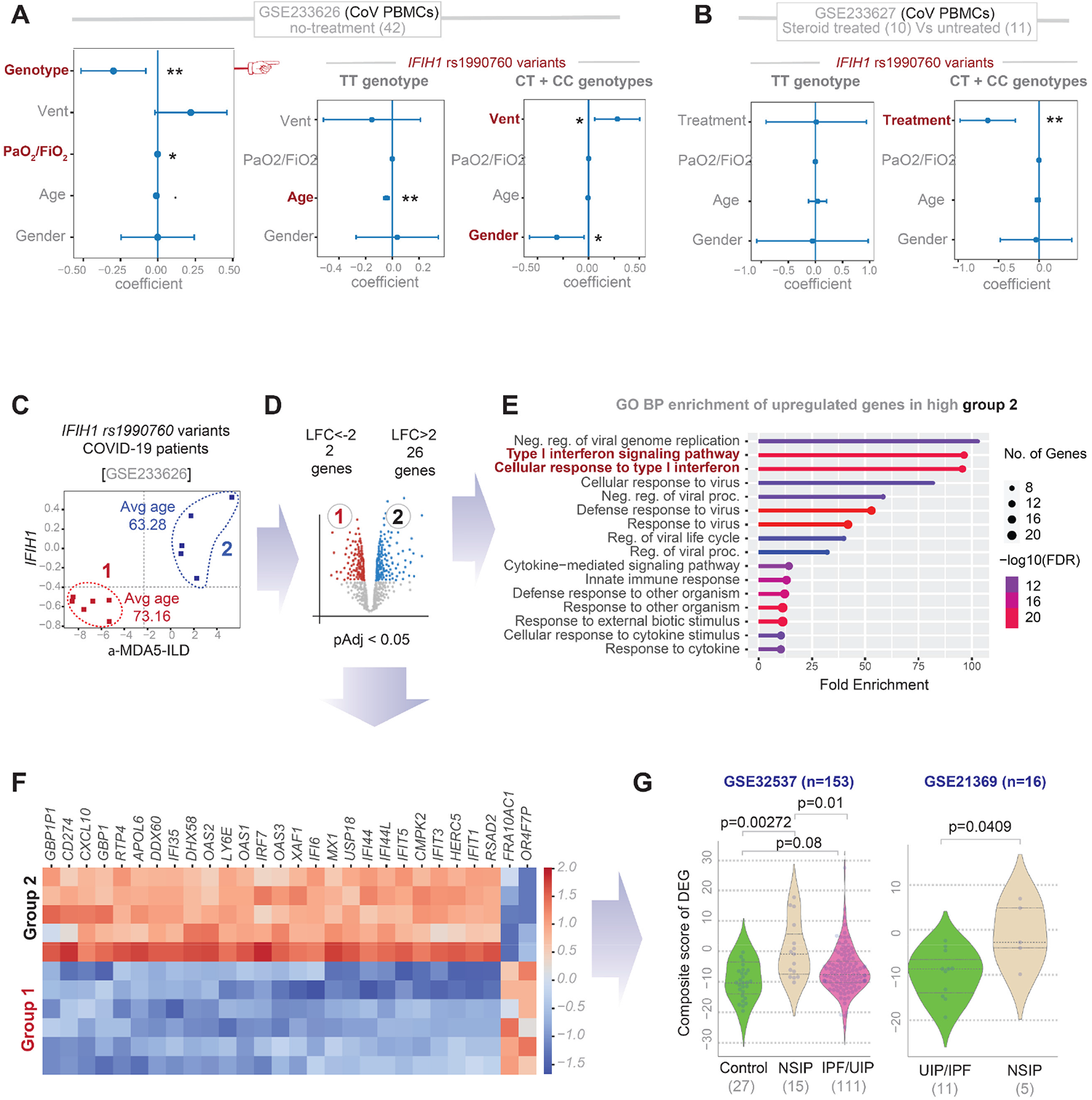
The rs1990760 TT variant of *IFIH1* offers an age-dependent protection against MDA5 surge. **A-B.** Multivariate analysis of *IFIH1* expression as a linear combination of all variables in the COVID-19 PBMC datasets GSE233626 (A) and GSE233627 (B). Coefficient of each variable (at the center) with 95% confidence intervals (as error bars) and the p values were illustrated in the bar plot. The p-value for each term tests the null hypothesis that the coefficient is equal to zero (no effect). Red = significant co-variates. **C-E.** Two distinct subgroups of COVID-19 patients with the rs1990760 TT genotype (groups 1 and 2 in the scatter plot in A) were assessed for differentially expressed genes (DEGs; B). Lollipop graph (C) displays the findings of a gene ontology (GO) analysis on the list of 26 genes upregulated in group 2. **F**. Heatmap displays DEGs (26 up-and 3 down-regulated; LogFC >2, pAdj 0.05) in group 2 PBMCs compared to group 1. **G**. Violin plots display the composite score of the DEGs (used as a gene signature) in two independent transcriptomic datasets of lung tissues from subjects with undefined (UIP) or non-specific (NSIP) interstitial pneumonitis and non-diseased controls.

A similar analysis on another independent dataset in which intervention was performed in the form of systemic corticosteroid treatment. Such treatment is an independent protective factor exclusively in the subjects with rs1990760 CT/CC variants, but not in those with the rs1990760 TT variant (**Figure 5B**). Taken together, these findings reveal a complex interplay between *IFIH1* genotype in which the rs1990760 TT variant offers age-dependent protection to the elderly. Among those who lack this protective variant, female gender and less severe disease increases the degree of *IFIH1* surge, whereas systemic therapy with steroids offers protection.

### The nature of the immunophenotype associated with the induction of MDA5 (*IFIH1*) transcript

We asked if *IFIH1* induction may be associated with an age-dependent immunophenotype that modulates the risk of progressive autoimmune ILD. We assessed the differentially expressed genes (DEGs) between the two distinct groups of patients within the rs1990760 TT variant, i.e., low-and high-inducers of the *IFIH1* transcript (**Figure 5C-D**). The *IFIH1*-high group induced 26 genes that are enriched for type 1 IFN signals and cellular responses to the same (**Figure 5E-F**). Upregulated genes are notable for markers of progressive autoimmune ILD, e.g., *CXCL10* ^45^, IFN-induced genes associated with systemic autoimmune rheumatic diseases (SARD) [*IFI44L*, *LY6E*, *OAS3*, *RSAD2* ^46^], adaptive immune hallmarks of MDA5+ DM [*IFI6, MX1, OAS2* ^42^] and *MX1* ^47^ (**Figure 5F**). These DEGs were significantly induced in autoimmune ILD (**Figure 5G**; non-specific interstitial pneumonitis, NSIP), compared to IPF (usual interstitial pneumonia, UIP). Similarly, when we analyzed the DEGs in lung epithelial cells that were treated with acute vs convalescent serum derived exosomes, we found that the Type 1-centric genes induced in the lung epithelium were significantly induced also in NSIP compared to IPF (**Supplementary Figure 2**).

## Discussion

Several COVID-19 era case reports or series of MDA5+ myositis or ILD have been reported in the UK and internationally either in the setting of infection or post-vaccination ^1–4,10–28^. Our study is the largest one to document the features and outcomes of this clinical syndrome, especially in 2021. Approximately 42% of our MDA5+ cases have thus far had progressive ILD, with a third of these proving fatal so far, in keeping with the known aggressive course of MDA5^+^-ILD ^48,49^. Our clinical epidemiologic observations, together with the transcriptomic analyses suggest that increased incidence of MDA5 autoimmunity and ILD that presented contemporaneously during COVID-19 could be due to an abberant type 1-centric IFN responses that are shared with autoimmune ILD, but not IPF, which plays out across diverse cell types leading to severe ILD (**Figure 6**).

**Figure 6.**
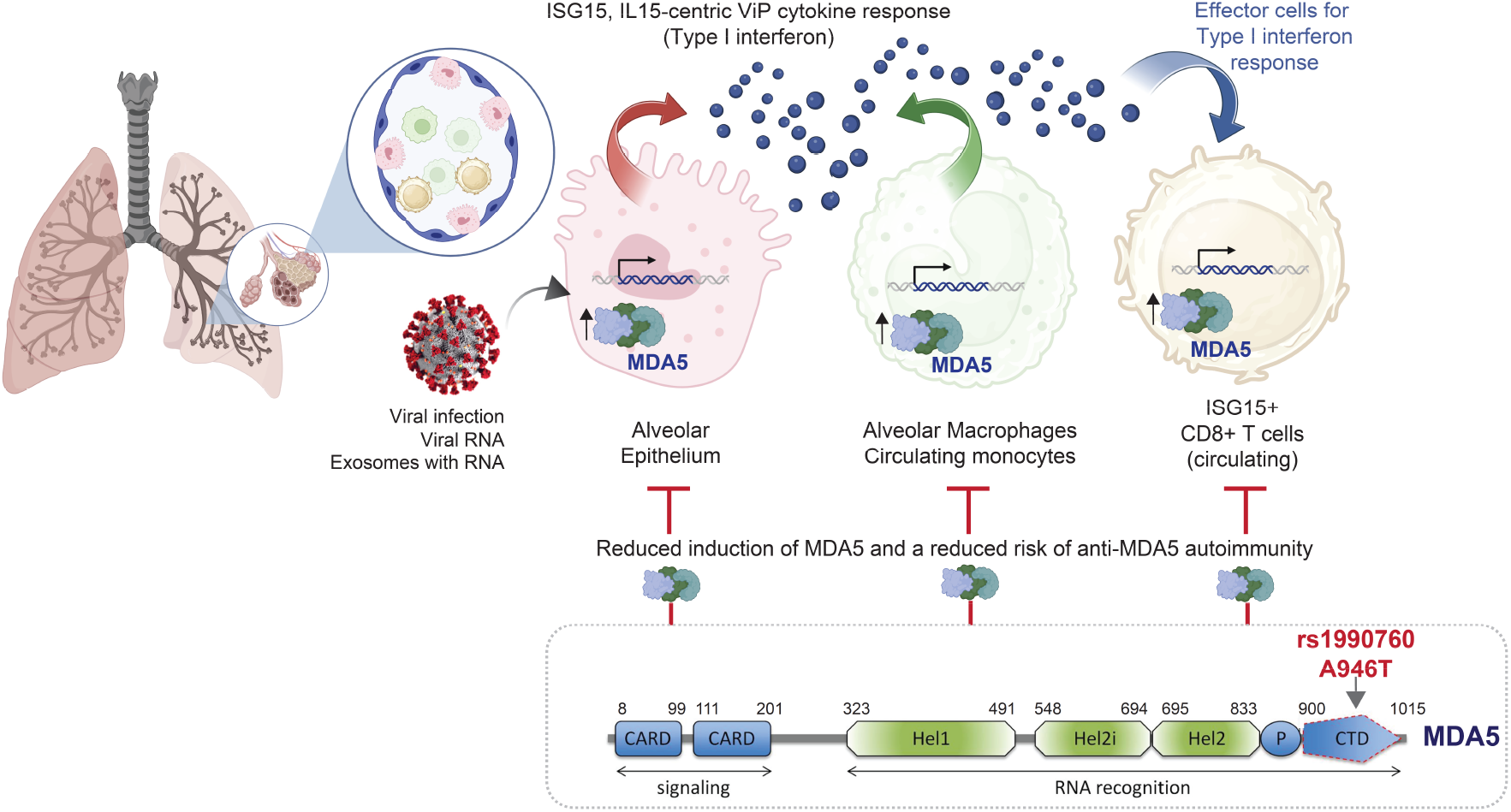
Summary and working model. Schematic summarizes major conclusions and a proposed working model. A type 1-centric interferon response to the same could serve as pathophysiologic driver of autoimmune ILD involving more than one cell type. From left to right (*<u>Top</u>*): (i) In the alveolar pneumocytes of COVID-19 lungs, MDA5 is induced and is associated with type 1 interferon response, AT2 senescence and stem cell dysfunction. MDA5 is induced also in lung epithelial cells upon exposure to exosome vesicles from patients with acute infection. (ii) In the PBMCs of COVID-19 patients MDA5 is induced in infected samples, and its degree of induction positively and tightly correlates with an IL-15 centric type 1 interferon response. (iii) In the PBMCs of COVID-19 patients, there is a concomitant induction of a signature for anti-MDA5 autoimmune ILD expressed in ISG15+ CD8+ T cells. Bottom panel shows the impact of a protective genotype of the *IFIH1* gene which inhibits a subset of patients from inducing MDA5 and thereby protects them from a surge of type 1 interferon storm.<colcnt=1>

Our observations, taken together with global reports of similar cases, leads us to propose the term *<u>M</u>*DA5-autoimmunity and *I*nterstitial *<u>P</u>*neumonitis Contemporaneous with the *<u>C</u>*OVID-19 Pandemic (MIP-C) (**Table 2**). Such an acronym has credence because of the distinct features that separate MIP-C from the syndrome of MDA5+ DM ^50^ including our population being predominantly Caucasian instead of the historically reported MDA5^+^-DM East Asian predilection and the lower rate of ILD that was evident in 42% of cases, at least thus far, to that historically reported in MDA5^+^-DM ^51–53^. Also the pathogenesis of MDA5^+^-DM is poorly understood but our work in 60 new cases and that from around the world ^2–4,15,18,22,54–67^ shows good evidence for a link to SARS-CoV-2 infection and vaccination and possibly both (**Figure 2**).

**Table 2:** Comparison between “classic” MDA5+ disease and MIP-C.

|  | Classic MDA5<sup>+</sup>-Disease | MIP-C |
| --- | --- | --- |
| **Age** | Adults<br>7% MDA5<sup>+</sup> among cases of juvenile dermatomyositis <sup>87,88</sup> | 4/60 cases (6.6%) children |
| **Gender** | Females 66% <sup>50</sup> | 36/60 cases (60%) females |
| **Ethnic background** | Asian | 32/60 cases (53%) white (British or any other “white” category)* |
| **Lung involvement** | Almost universal in people of Asian descent.<br>Pulmonary involvement reported between 39% and 73% elsewhere globally (Brazil, Italy, Spain, North America) <sup>53</sup> | 25/60 cases (41.6 thus far) |
| **Interstitial lung disease prognosis** | Poor, frequently fatal in adults | Fatalities less common, though progressive pulmonary function deterioration frequent (8/60 -13.3%) |
| **Isolated, Non-Pulmonary Disease** | Uncommon | 35/60 cases (58.3%) experienced manifestations of connective tissue diseases (18/60 cutaneous rash; 20/60 Raynaud’s phenomenon; 5/60 “mechanic’s hands”, some of the 35 have more than one combined) |
| **Associated antibody positivity** |  | About two third cases have associated antibody positivity (36/60), being anti Ro 52 (13/60) and Anti SAE1 (12/60) the most common. |

In the Yorkshire region of UK, the estimate for population classified as “white” (any “white” category) was 85.4%, according to 2021 UK census. Such percentage reflects the difference in prevalence recorded for the whole England and Wales population (81.7%) at the time of 2021 UK census. \*14/60 patients had no ethnicity available.

The MIP-C phenotype, somewhat akin to MIS-C in children, quite often had no history of confirmed SARS-CoV-2 infection. Given that nearly 42% of new cases were not vaccinated prior to MDA5+ disease, it suggests that milder COVID-19 disease, either overt, or covert (i.e., asymptomatic infection or incidental exposure) may be sufficient to cause MDA5 autoimmunity. Given the peak of MDA5 positivity testing followed the peak of COVID-19 cases in 2021, and coincided with the peak of vaccination, these findings suggest an immune reaction or autoimmunity against MDA5 upon SARS-CoV-2 and/or vaccine exposure; it could represent novel immunogenicity in non-immune subjects upon RNA engagement with MDA5, causing a surge of cytokine response, and then the triggering of an autoimmune disease. The development of herd immunity and less respiratory exposure to to SARS-CoV2 could theoretically contribute to the milder phenotype at the population level in our proposed MIP-C entity.

As for how COVID-19 vaccine may give rise to such immunogenicity, a recent study by Li et al., has shed some light ^68^. The authors showed that in the lymph nodes (LNs), modified RNA sensed by MDA-5 results in the production of type I interferons (IFNs); the latter induce antigen-specific CD8+ T cell responses ^68^. This conclusion was derived after the authors systematically evaluated the immunogenicity response to BNT162b2 LNP-mRNA against COVID-19 in numerous murine models lacking RNA-sensing pattern recognition receptors [Toll-like receptors 2, 3, 4, 5 and 7 and other inflammosome and necroptosis/pyroptosis pathways] where only MDA-5 was deemed important for type I interferon responses and for antigen-specific CD8^+^ T cell responses ^68^. Because RNA can be recognized by MDA5 in a sequence and structure-dependent manner ^69^, the resultant activation of the innate immune system is believed to be cell, tissue and context specific. Our finding incriminate MDA5 protein activation, whether linked to natural infection, or vaccination or potentially both as a trigger for MIP-C and that MDA5-mediated sensing (and mounting of an immunophenotype that is comprised of type 1 interferonopathy and antigen-specific CD8^+^ T cell responses; elaborated below) is a distinct trigger in MIP-C.

There are four noteworthy findings that inform how we recognize and/or manage MIP-C in the aftermath of COVID-19. First, that the viral sensor *IFIH1*/MDA5 is induced in COVID-19 has been reported exhaustively ^9,70–77^. We found that the severity of COVID-19 may dictate the risk of progression to ILDs of distinct immunopathogenesis: Milder disease induced *IFIH1* and risk signatures for MDA5-autoimmunity; however, severe disease with diffuse alveolar damage in the setting of acute respiratory distress syndrome (ARDS) induced risk signatures for alveolar dysfunction that are pathognomonic of IPF, consistent with prior claims ^30^.

Second, our finding that the degree of *IFIH1* induction is strongly associated with the degree of induction of a type 1 IFN signature hat is quite specific for being IL-15-centric [ViP signature ^35^] is noteworthy. This finding is in keeping with prior work showing the importance of this IL-15 in rapidly progressing ILD in the setting of MDA5 autoimmunity and amyopathic DM ^78–80^. Given the extensive literature on the role of the IL15/IL-15RA axis in the development of autoimmunity [reviewed in ^81^], and more specifically its role in triggering the activation of CD8+ T cells to drive such autoimmunity ^82–85^,

Third, the recognition of MIP-C as a syndrome where less than half of cases get severe progressive ILD is relevant for therapy selection including Janus kinase (JAK) inhibitors, such as tofacitinib ^86^ as many cases did not progress, at least in the first two years of MDA5+ status. Fourth, we show that the rs1990760 (p.Ala946Thr) *IFIH1* variant displays, what is likely to be an age-dependent protection ^74^, to a subgroup of patients; these patients show a lesser induction of *IFIH1,* a blunted type 1 IFN storm, and a reduced signature of circulating *ISG15*^+^CD8^+^T cells which was previously found to predict poor one-year survival in MDA5^+^DM patients ^42^.

Our study has some limitations, including the retrospective nature of the clinical data collection and uncertainties around the confirmation of COVID19 infection status (most patients were not systematicaly tested) and could be infected but asymptomatic. Furthermore, we have no data on asymptomatic infection or prolonged carriage status as potential factors in some of these cases; neither did we have patient-derived samples to analyse transcriptomic datasets from our cohort. We also do not delineate how autoimmunity arises; given that MDA5 is a key RNA receptor in the lung parenchymal and immune cells it is tempting to speculate that MDA5 and nucleic acid as an antigen and associated bound adjuvant could contribute to triggering autoimmunity. A clear mechanism for the vascular basis for the DM and PSS lesions is yet to emerge. Regardless, we have shown in numerous independent cohorts that the degree of induction of *IFIH1* (MDA5) is tightly correlated with the degree of induction of type 1 interferons and a gene signature for risk of progressive MDA5^+^ILD.

In conclusion, in this work we report a remarkable rise in MDA5+ disease in the Yorkshire region that, given the overall epidemiology, we have termed MIP-C. We provide transcriptome derived insights that point to a plausible and potentially causal link between the surge in anti-MDA5-positivity, autoimmune ILD and COVID-19, but not IPF. These findings warrant further studies, preferably through multi-centre efforts and across nations, to begin to recognize and better appreciate the potential global clinical burden of interstitial pneumonitis and ILD in the aftermath of the COVID-19 pandemic.

## Acknowledgements

We thank Guillermo M Albaiceta (Hospital Universitario Central de Asturias, Oviedo, Spain) for access to demographic metadata on published datasets and Debashis Sahoo (UC San Diego) for access to computational tools at the Center for Precision Computational Systems Network (PreCSN). This work was supported in part by the National Institute for Health Research (NIHR) Leeds Biomedical Research Centre (BRC), and in part by the National Institutes for Health (NIH) grant R01-AI155696 and pilot awards from the UC Office of the President (UCOP)-RGPO (R00RG2628, R00RG2642 and R01RG3780) to P.G. S.S was supported in part by R01-AI141630 (to P.G) and in part through funds from the American Association of Immunologists (AAI) Intersect Fellowship Program for Computational Scientists and Immunologists. The views expressed are those of the author(s) and not influenced by the study funders, the NHS, the NIHR or the Department of Health.

## Data Availability

All data produced in the present work are contained in the manuscript

## Author Contributions

SS conducted all the statistical, mathematical, computational, or other formal techniques; ST and EM assistated with dataset processing and curation; KI PD and GDM created all Tables for visualization and data presentation; SS and PG created all figures for visualization and data presentation; DMG conceptualized and supervised all clinical aspects of this study; PG conceptualized and supervised all computational aspects of this study; DM and PG jointly administered the project and secured funding; KI, PD, GDM, DMG and PG wrote initial draft; all authors edited the manuscript and approved its final version.

