## Supplementary Online Materials for "MDA5-autoimmunity and Interstitial Pneumonitis Contemporaneous with the COVID-19 Pandemic (MIP-C)"

[1]Leeds Teaching Hospitals NHS Trust, Rheumatology Department, Leeds, United Kingdom. [2]Department of Cellular and Molecular Medicine, School of Medicine, University of California San Diego, La Jolla, CA, 92093, USA. [3]Mid Yorkshire Teaching NHS Trust, Rheumatology, Wakefield, United Kingdom [4]University of Leeds, Leeds Institute of Rheumatic and Musculoskeletal Medicine, Leeds, United Kingdom. [5]NIHR Leeds Biomedical Research Centre, Leeds Teaching Hospitals NHS Trust, Leeds, United Kingdom. [6] Department of Computer Science and Engineering, Jacob's School of Engineering, University of California San Diego, La Jolla, CA. 92093, USA. [7]Leeds Teaching Hospitals NHS Trust, Pathology, Leeds, United Kingdom, [8]University of Leeds, Immunology, Leeds, United Kingdom. [9]Bradford Teaching Hospitals NHS Foundation Trust, Rheumatology, Bradford, United Kingdom. [10]Harrogate and District NHS Foundation Trust, Rheumatology, Harrogate, United Kingdom. [11]Airedale NHS Foundation Trust, Rheumatology, Steeton with Eastburn, United Kingdom. [12]Calderdale and Huddersfield NHS Foundation Trust, Rheumatology, Huddersfield and Halifax, United Kingdom. [13]Department of Medicine, School of Medicine, and Veterans Affairs Medical Center, University of University of California San Diego, La Jolla, CA. 92093, USA.

‡**Corresponding Authors**:
Dennis McGonagle PhD FRCPI
Leeds Teaching Hospitals NHS Trust, Rheumatology Department, Leeds, United Kingdom | University of Leeds, Leeds Institute of Rheumatic and Musculoskeletal Medicine, Leeds, United Kingdom |

Pradipta Ghosh, M.D.
Professor, Departments of Medicine and Cellular and Molecular Medicine, University of California San Diego; 9500 Gilman Drive (MC 0651), George E. Palade Bldg, Rm 232, 239; La Jolla, CA 92093, USA | Phone: 858-822-7633 |

#### INVENTORY OF SUPPLEMENTARY MATERIALS

- Supplementary Figures (2)
- Supplemental Tables (2)
- Supplementary Information (2). These are uploaded separately as excel datasheets

### Supplementary figures

#### Supplementary figure 1

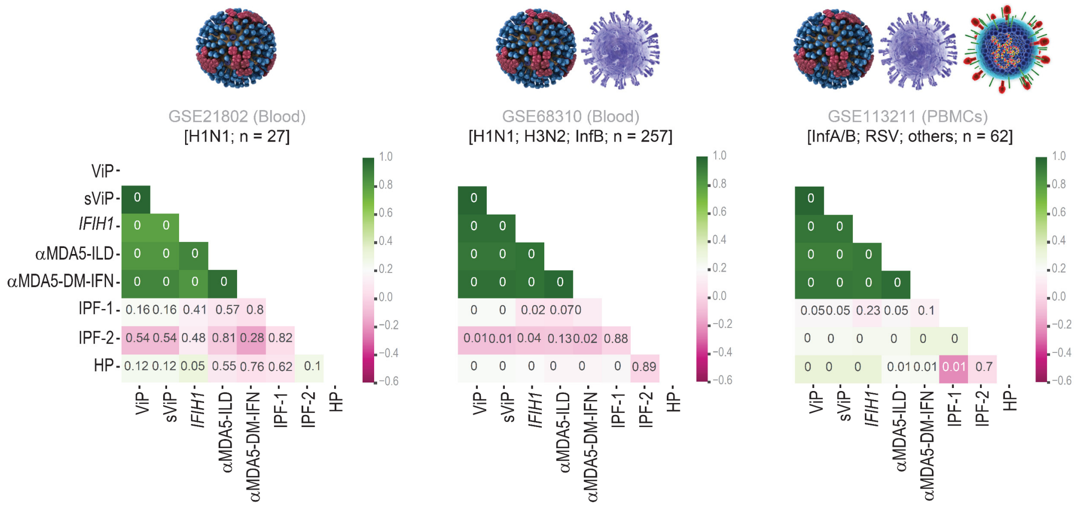


**Supplementary Figure 1: [Related to Figure 4]**

**Induction of *IFIH1* in respiratory viral pandemics correlates with the induction of Type 1-centric cytokine storm and α-MDA5-ILD risk signatures in whole blood and PBMCs.** Graphical representation of a correlation matrix representing the pair-wise correlation between multiple variables, i.e., composite scores of different gene signatures elaborated in **Figure 4A**. The colour key spans from -1 (magenta) to 1 (green), indicating both strength and direction of correlation. Numbers within the heatmap indicate statistical significance (only significant *p values* are displayed).

#### Supplementary figure 2

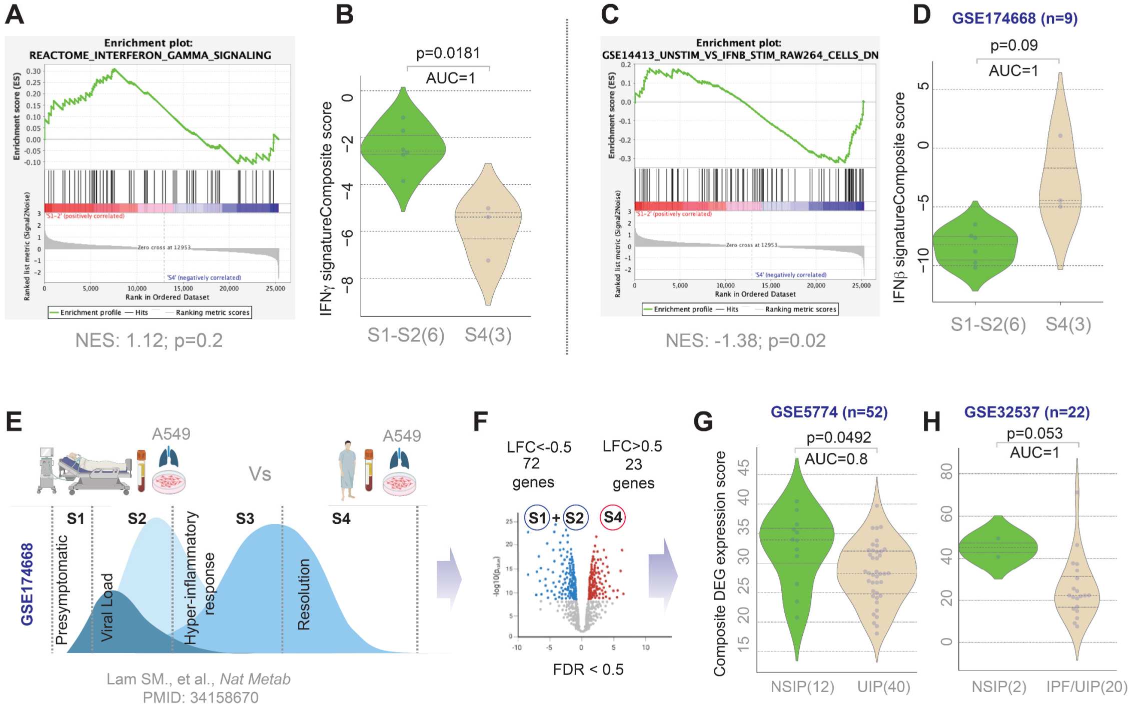


**Supplementary Figure 2**: **[Related to Figure 6]**

**Serum derived exosomes from convalescent patients with CoV induce a Type 1-centric immunophenotype that is induced also in the lung tissues from non-specific interstitial pneumonitis (NSIP).**

**A-D.** Gene Set Enrichment Analyses (GSEA pre-ranked analysis) of Type 1 and 2 interferon response derived from MSigDB demonstrate the differential activation of these interferons in lung epithelial cells exposed to exosome vesicles from the serum of early (A-B) and late (C-D) stages of COVID-19 infection.

**E-F.** Schematic in panel E shows the workflow for the derivation of differentially expressed genes (DEGs; panel F) in lung epithelial cells exposed to exposed to exosome vesicles from the serum of early (S1-2) and late (S4) stages of COVID-19 infection.

**G.** Violin plots display the composite score of the DEGs (used as a gene signature) in two independent transcriptomic datasets of lung tissues from subjects with undefined (UIP) or non-specific (NSIP) interstitial pneumonitis and non-diseased controls.

#### Supplemental Table 1- MDA5 Related ILD Patterns

| Gender, age (age range) | ethnicity | past history of autoimmunity | COVID19 vaccination (first dose) | COVID19 infection status | Non-clinical ILD features | Baseline CK | ANA IIF | Other myositis Ab | Imaging | Treatment | Treatment outcome |
|---|---|---|---|---|---|---|---|---|---|---|---|
| Male, (45-49) | white-british | nil | 7 months after, no flare | not known | proximal muscle weakness, inflammatory arthritis, cutaneous features (ex. Gottron's) | 304 | negative | anti-PL7+ | Chest CT with pulmonary fibrosis and ground glass in the bases, pattern consistent with ILD | MTX (2020, for arthritis), IVIg (during septic arthritis), MMF (nonresponse), cyclophosphamide pulses, HCQ (stopped due to retinal damage) | progressive lung disease |
| Male, (85-89) | white-british | nil | 3 months after, no flare | 10 months after, no flare | none | 178 | negative | anti-SAE1 | Chest CT with NSIP pulmonary fibrosis picture | nintenamib | progressive lung disease |
| Female, (50-54) | white-british | nil | 2 months after, no flare | not known | heliotrop rash, shwal, gottren's, synovitis | 92 | positive | none | Chest CT initially normal but after a year central mid and upper zone ill defined areas of ground glass opacification, no pulmonary fibrosis | MTX, prednisolone, nifedipine | stable lung disease |
| Female, (65-69) | n/a | nil | 4 months before | 1 year and 4 months after, no flare | raynaud | 149 | positive | none | Chest CT with NSIP pattern pulmonary fibrosis | Rituximab, MMF, cyclophosphamide pulse then MMF again | died (ILD) |
| Female, (60-64) | white-british | Hypothyroidism? | 2 months after, no flare | not known | none (possibly synovitis) | 71 | negative | anti-TIFFgamma | Chest-CT with co-existing pulmonary fibrosis and emphysema (smoker) | nintenamib | progressive lung disease |
| Male, (40-44) | any other Asian | nil | not vaccinated | not known | synovitis, possibly gottron's, possibly heliotropic, slighly proximal weakness in upper limbs | 47 | negative | anti-Ro52 | CT angiogram pulmonary - negative for PE, but bilateral ground glass changes | 3x doses IV methylprednisolone and then oral prednisolone, IV cyclophosphamide, 5 days of IVIG, tacrolimus and HCL, Rituximab | died (ILD) |
| Male, (60-64) | white-british | nil | 2 weeks before | not known | none | 33 | positive | anti-SAE1 | Most recent chest CT with progressive interstitial lung changes with new ground glass opacification and | methylprednisolone IV | died (ILD) |

| | | | | | | | | | | | |
|---|---|---|---|---|---|---|---|---|---|---|---|
| | | | | | | | | | pulmonary fibrosis, new bilateral effusions | | |
| Female, (30-34) | white-british | nil | 11 months before | not known | mechanic hands | 52 | positive | anti-SAE1 | Most recent chest CT when compared to exam performed six months before, there were new areas of ground-glass changes/consolidation (both centrally and peripherally) in the upper lobes bilaterally and middle lobe. | methylprednisolone IV, cyclophosphamide, Rituximab, MMF | died (ILD) |
| Female, (55-59) | "other ethnic group" | nil | 3 months before | 1 year and 8 months after, no flare | mechanic hands, SLE overlap | 105 | positive | Anti-SAE1, Anti-SRP | Chest CT with upper lobe pulmonary fibrosis and ground glass appearance compatible with ILD | long term steroids, cyclophosphamide, rituximab, azathioprine (intolerant, stopped), MMF, IVIg | stable lung disease |
| Female, (70-74) | white-british | nil | 1 day after | not known | none | 29 | positive | none | Chest CT with significant amount of ground glass change with subpleural sparing, compatible with NSIP | none (waiting on tests to differentiate from hypersensitivity) | died (ILD) |
| Female, (60-64) | white-british | Grave's disease | 5 months before | 10 months before, flared | raynaud | 78 | negative | none | Chest CT with upper lobe fibrosis with honeycombing | none | stable lung disease |
| Female, (75-79) | white-british | hypothyroidism? | 5 months after, flared | not known | sicca symptoms | 50 | negative | Anti-SAE1 | Chest CT with UIP fibrosis | none | stable lung disease |
| Male, (35-39) | Pakistani | nil | not vaccinated | not known | cutaneous rash (gottron's papulles, facial malar/abdominal rash) | 299 | positive | none | Chest CT with pneumopericardium and mediastinum with multifocal consolidation and ground glass changes alongside with traction bronchiectasis | methylprednisolone IV, IVIg, cyclophosphamide | died (ILD) |
| Female, (10-14) | Pakistani | nil | not vaccinated | 1 year and 3 months before | inflamatory arthritis, sclerodermatous skin and calcinosis | 105 | positive | anti-Ro52 | Chest CT with some pulmonary fibrosis | azathioprine, MMF, Rituximab, IVIg, sodium thiosulphate, HCQ, presnisolone | stable lung disease |
| Female, (50-54) | n/a | nil | not vaccinated | not known | none | n/a | negative | anti-SAE1 | Chest CT with round glass changes with traction of the airways | none (waiting on tests) | died (ILD) |

| | | | | | | | | | | | |
|---|---|---|---|---|---|---|---|---|---|---|---|
| | | | | | | | | | just patchy and geographical and worse at the bases (not NSIP pattern, could be CTD-ILD) | | |
| Female, (50-54) | not given | nil | 3 months before | 2 months after, flared | periorbital rash, also in upper arm and wrists | n/a | negative | anti-PL7+, anti-Ro52, anti-PScl100 | Chest CT with ground glass changes compatible with ILD | cyclophosphamide, steroids, MMF | stable lung disease |
| Male, (70-74) | white-british | nil | 8 months before | not known | none | 47 | negative | anti-SAE1 | Chest CT with mild fibrosis, interstistial changes and subtle sub-pleural abnormalities | none | stable lung disease |
| Male, (80-84) | not given | nil | 1 year and 5 months before | not known | none | 68 | positive | none | Chest CT with bilateral mainly peripheral and basal fibrosis with ground glass changes and bronchiectasis | none | stable lung disease |
| Male, (85-89) | white-british | nil | 1 year and 8 months before | not known | none | 63 | negative | anti-PL2, anti-SRP, anti-Mi-2-alpha | Chest CT showing fibrosis, emphysema and true lower lobe bronchiectasis | none | stable lung disease |
| Male, (50-54) | Pakistani | nil | 6 months before | not known | none | 136 | positive | none | Chest CT showing extensive patchy bilateral ground glass changes, bronchiectasis | steroids, to start nintedanib | progressive lung disease |
| Male, (70-74) | white-british | nil | 1 year before | 1 year before | none | 116 | positive | none | Chest CT showing pulmonary fibrosis and nodules | none | stable lung disease |
| Male, (55-59) | n/a | nil | 1 week before | not known | hemoptysis (self-resolved) | n/a | positive | anti-Ro52 | n/a | n/a | n/a |
| Female, (70-74) | n/a | nil | 1 year and 8 months before | not known | raynaud, synovitis, rashes | n/a | positive | anti-PMScl100 and anti-PMScl70 | Chest CT with usual UIP | MTX | stable lung disease |
| Male, (30-34) | n/a | nil | not vaccinated | 5 days before | mild synovitis, stiffness and weakness with power reduced in lower limbs and some | 27 | positive | none | Chest CT scan 2 months prior to admission demonstrated bilateral peripheral ground glass change in both lungs. In admission proggression of changes compatible | steroids, cyclophosphamide, rituximab, tofacitinib, plasma exchange | intial partial response but than progresssive disease and died |

| | | | | | | | | | | | |
|---|---|---|---|---|---|---|---|---|---|---|---|
| | | | | | breathlessness, rash | | | | with rapid proggressive ILD. | | |
| Male, (70-74) | n/a | nil | 1 year and 10 months before | not known | none | 206 | positive | none | CT with non-typical ILD findings | none | stable lung disease |

**ILD = Interstitial Lung Disease; CK = Creatin-Kinase; ANA IIF = Anti-Nuclear Antibodies Indirect ImmunoFluorescence; N/A = Not Available; CT = Computer Tomography scan; CTD = connective tissue disease; UIP = Usual Interstitial Pneumonia IV = Intra-Venous; MMF = Mophetyl Mychophenolate; MTX = Methotrexate, HCQ = HydroxyChloroQuine; IVIg = IV Immunoglobulins; not known for COVID-19 infection status considered when not tested**

#### Supplemental Table 2: Characteristics of Non-ILD patients

| Gender, age (age range) | ethnicity | past history of autoimmunity | COVID-19 vaccine status | COVID-19 infection before (how long) | Non-clinical ILD features | Baseline CK | ANA IIF | Other myositis Ab | Relevant Imaging | Treatment | Treatment outcome |
|---|---|---|---|---|---|---|---|---|---|---|---|
| Female, (35-39) | any other Asian | nil | 1 month after, no flare | not known | Symmetrical proximal myositis, gottrons's papules, photosensitivity, inflammatory arthritis | 123 | positive | none | proximal myositis in femur MRI | prednisolone, MTX | partial response |
| Male, (45-49) | white-British | nil | 1 year and 9 months after, cutaneous flare | 11 months after, no flare | overlap connective tissue disease with dermatomyositis and systemic sclerosis (Raynaud's, synovitis, cutaneous ulcers, dermatomyositis rash and mouth ulcer) | 89 | negative | none, but SCL70 | n/a | rituximab, HCQ, MMF, PPI | improvement |
| Male, (50-54) | any other Asian | nil | 1 year after, no flare | 11 months before | right tight myositis | 33 | negative | AntiRO52, antiOJ, anti-PL7, Anti-PM-Scl75, anti-KU, Anti SAE1, anti NXP2, anti-TIF1, anti MI-2 alpha | MRI and US thigh normal muscle appearance | none | total improvement |
| Female, (80-84) | white-British | nil | 1 week after, flare? Trigger? | not known | Raynaud, non-myositis | 126 | positive | none | none | nifedipine | improvement |
| Female, (50-54) | white-Irish | nil | 5 months before | not known | proximal myopathy | 280 | positive | none | Thigh MRI compatible with myositis | MMF, prednisolone, rituximab | improvement |
| Male, (45-49) | white-British | sarcoidosis? | 2 months after, no flare | not known | Raynaud, periungal changes, non-myositis | 481 | negative | anti-SAE1 | none | none | stable |

| | | | | | | | | | | | |
|---|---|---|---|---|---|---|---|---|---|---|---|
| Female, (55-59) | white-British | nil | 3 months before | not known | Raynaud, synovitis | 66 | positive | anti-Ro52 | none | nifedipine | progression |
| Female, (75-79) | white-British | nil | 3 months before | 2 months before | neck pain, progressive breathlessness | 220 | positive | anti-PMScl100 | Chest CT with no ILD, no evidence of myositis in neck MRI | none | stable |
| Female, (25-29) | N/A | nil | 6 days before | not known | Raynaud | 62 | positive | none | none | none | stable |
| Female, (40-44) | black-Caribbean | nil | not vaccinated | not known | inflammatory arthritis, Raynaud, two miscarriages (APL negatives) | 63 | negative | anti-Ro52, anti-PL7, anti PScl100 | none | MTX, steroids, HCQ, azathioprine | no response |
| Male, (70-74) | N/A | nil | 1 year and 6 months before | not known | haemoptysis and haematuria | n/a | positive | anti-PL7, anti SAE1, anti-ku, anti NXP2, anti-Ro52 | Chest CT with no ILD | none | improvement |
| Female, (35-39) | Pakistani | SLE | 2 years and 5 months after, no flare | 2 years and 10 months after, no flare | nail fold abnormalities, leukopenia, panniculitis, malar rash, heliotrope rash, digital maculopathy, myositis | 33 | negative | none | signs of myositis in MRI of proximal inferior limbs | prednisolone, HCQ, MMF and 1 cycle rituximab | partial response |
| Male, (5-9) | white-British | nil | not vaccinated | not known | sore gums, mouth ulcers, gottrons's papules, polyarthritis, fever, mild weakness on legs | 211 | negative | anti-Ro52 | MRI thighs with widespread patchy muscle oedema | IV methylprednisolone, MMF, MTX | no response |
| Female, (70-74) | white-British | amyopathic dermatomyositis anti-MDA5- but ntiRo52 and Ant-iKu + | 5 days after | 2 weeks before, Trigger? | calcinosis, dermatomyositis cutaneous changes (not specified) | 174 | positive | anti-Ro52, anti-NPX2 | MRI thigh normal, pulmonary function test normal | oral steroids, MTX | stable |
| Female, (10-14) | black-African | Sjogren syndrome | not vaccinated | not known | synovitis, sicca syndrome, weak proximal limbs | 115 | positive | anti-Ro52 (was + in 2018, MDA5 was -) | MRI compatible with myositis | MTX, MMP, HCQ, rituximab | partial response |
| Female, (10-14) | not given | nil | not vaccinated | not known | malaise, léthargie, joint pains, Raynaud | 25 | negative | anti-PMScl75, | none | none | improvement |

| | | | | | | | | | | | |
|---|---|---|---|---|---|---|---|---|---|---|---|
| Male, (85-89) | white-British | nil | 8 months before | not known | dysphagia | n/a | positive | anti-Mi2-beta, anti-Mi-2-alpha | n/a | n/a | n/a |
| Male, (50-54) | white-British | nil | 7 months before | not known | hyperkeratosis | 38 | negative | none | n/a | n/a | died |
| Female, (35-39) | white-black African | nil | 4 months before | 7 months before | Raynaud's phenomenon with Dermatomyositis like skin rash, mechanic hands, inflammatory joint pain, chronic fatigue | 76 | positive | none | none | methylphenidate | stable |
| Female, (35-39) | white-British | lupus | 7 months before | not known | Periungal inflammation, proximal weakness | 221 | positive | anti-Ro52 | n/a | MTX, HCQ, thalidomide, rituximab, iloprost, tacrolimuls | no response |
| Female, (65-69) | white-British | nil | 8 months before | 10 months after, no flare | Inflammatory arthritis, clubbing, oesophageal dysmotility, Raynaud | 29 | negative | none | Chest CT with no evidence of ILD, achalasia on chest CT | Rituximab, MTX, prednisolone, HCQ | no response |
| Male, (65-69) | white-British | nil | 3 months after, no flare | 1 month after, flared | Mild proximal weakness, inflammatory arthritis, lymphopaenia | 350 | negative | anti-SAE1 | none | MTX, prednisolone, nifedipine | improvement |
| Female, (40-44) | white-British | nil | 4 months before | not known | Raynaud | n/a | positive | none | n/a | n/a | n/a |
| Male, (45-49) | white-British | nil | 1 month before | not known | inflammatory arthritis and weight loss | n/a | positive | anti-SAE1 | n/a | n/a | n/a |
| Female, (35-39) | N/A | nil | not vaccinated | not known | Raynaud | n/a | negative | none | n/a | n/a | n/a |
| Female, (60-64) | N/A | nil | 1 year and 4 months before | not known | Raynaud | 76 | positive | anti-PMScl75, anti-NXP2, | none | none | stable |
| Female, (70-74) | any other Asian | Hypothyroidism? | 1 year and 4 months before | not known | Myositis, myocardial involvement, toe | 36 | negative | anti-Mi2-alpha | echo EF < 30%, MRI with myositis features (thigh) | prednisolone, MMF, rituximab | stable |

| | | | | | | | | | | | |
|---|---|---|---|---|---|---|---|---|---|---|---|
| | | | | | ulceration and Raynaud | | | | | | |
| Female, (60-64) | white-British | nil | 1 year and 4 months before | not known | Raynaud | 388 | positive | antiPL7 | normal EMG, normal chest CT, normal MRI thigh | none | improvement |
| Female, (25-29) | "other ethnic group" | crohn's disease, HS | not vaccinated | not known | Raynaud, puffy fingers, pitting scars | 81 | positive | anti-PMScl75 | early scleroderma on capillaroscopy, normal pulmonary function | nifedipine | progression |
| Female, (45-49) | white-British | nil | 1 year and 6 months before | not known | Raynaud | 121 | positive | none | US of shoulder with degenerative changes | none | stable |
| Female, (55-59) | white-British | nil | 4 months before | not known | MCTD symptoms | n/a | positive | anti-mibeta2, anti-PMScl75 | n/a | n/a | n/a |
| Male, (55-59) | n/a | nil | 1 year and 9 months before | not known | proximal myopathy, rashes, sicca syndrome, synovitis, Raynaud. | normal | positive | anti-RS, Anti-OJ, Anti-PL7, anti-EJ, Anti-Ku, Anti-NXP2, Anti-MRIFgamma2, Anti-Ro52 | n/a | none | stable |
| Female, (60-64) | black-Caribbean | nil | 7 months before | not known | myopathy, transient raised CK | 1715 | positive | none | no fatty atrophy or active myositis on imagining | observation/none | improvement |
| Male, (60-64) | white-British | nil | 1 month before | not known | myositis | 11000 | negative | none | myositis on MRI thighs biopsy suggestive of statin myositis HMGCoA Ab+ | prednisolone and MTX | improvement |
| Female, (80-84) | "other ethnic group" | PMR | 8 months before | not known | myositis | 22000 | positive | OJ Ab, EJ Ab, MPL122, PL-7, Ku,MTIFG2 | CT with no ILD | none | improvement |
| Female, (75-79) | white-Irish | nil | 10 months before | not known | myositis and synovitis | 55 | negative | none | CT with no ILD | prednisone | improvement |

**ILD = Interstitial Lung Disease; CK = Creatin-Kinase; ANA IIF = Anti-Nuclear Antibodies Indirect ImmunoFluorescence; N/A = Not Available; CT = Computer Tomography scan; MRI = magnetic resonance imaging; US (ultrasound); IV = Intra-Venous; MMF = Mophetyl Mychophenolate; MTX = Methotrexate, HCQ = HydroxyChloroQuine; PPI = proton pump inhibitors; not known for COVID-19 infection status considered when not tested**
